# Significant Production of Ozone from Germicidal UV Lights at 222 nm

**DOI:** 10.1101/2023.05.13.23289946

**Authors:** Zhe Peng, Douglas A. Day, Guy Symonds, Olivia Jenks, Harald Stark, Anne V. Handschy, Joost de Gouw, Jose L. Jimenez

## Abstract

Lamps emitting at 222 nm have attracted recent interest for germicidal ultraviolet disinfection (“GUV222”). Their impact on indoor air quality is considered negligible. In this study, ozone formation is observed for eight different lamps from five manufacturers, in amounts an order-of-magnitude larger than previous reports. Most lamps produce O_3_ in amounts close to the first-principles calculation, with e.g. a generation rate of 22 ppb h^-1^ for Ushio B1 modules in a 21 m^3^ chamber. Much more O_3_ is produced by lamps when optical filters were removed for tests, and by an undesired internal electrical discharge. A test in an office shows an increase of ∼6.5 ppb during lamp-on periods, consistent with a simple model with the O_3_ generation rate, ventilation and O_3_ losses. We demonstrate the use of a photolytic tracer to quantify the averaged GUV222 fluence rate in a room. Low-cost electrochemical O_3_ sensors were not useful below 100 ppb. Formation of O_3_ increases indoor particulate matter (PM), which is ∼10-30 times more deadly than O_3_ per unit mass, and which is ignored when only considering O_3_ threshold limit values. To limit GUV222-created indoor pollution, lower fluence rates should be used if possible, especially under low-ventilation conditions.

**TOC graphic:** 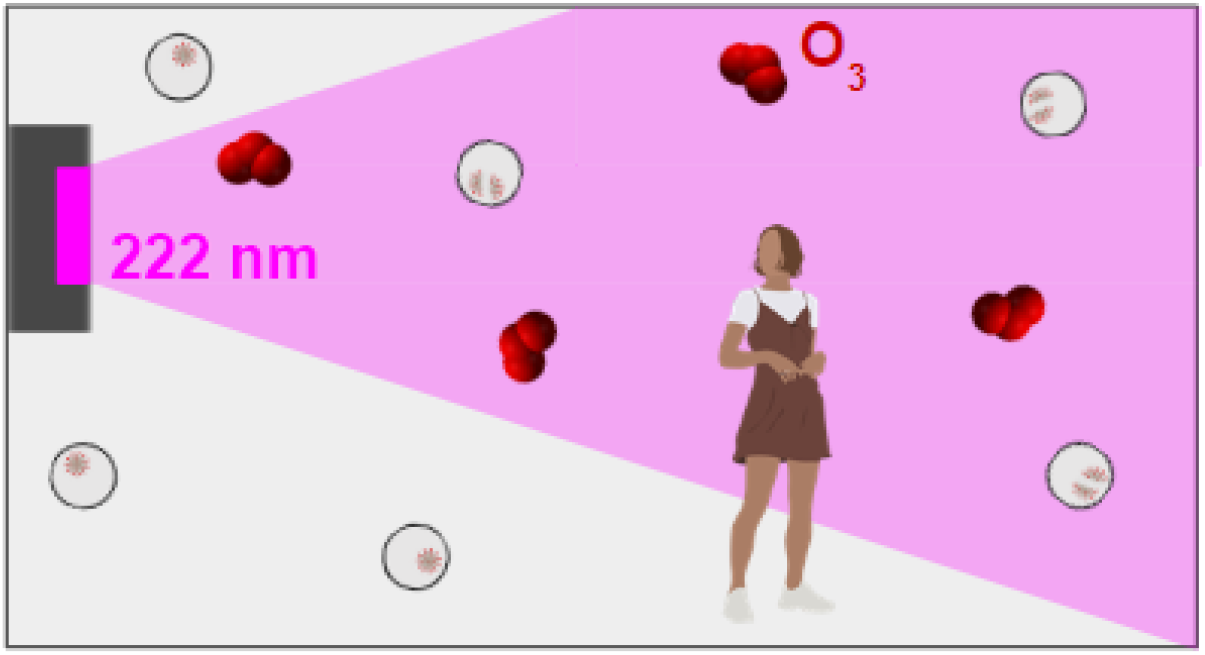

## 1. Introduction

Germicidal ultraviolet (GUV) disinfection has been used for a century to inactivate airborne pathogens, i.e. those that infect via inhalation of pathogen-containing aerosols that float in the air.^1–3^ Despite some early interest in widespread deployment (e.g. a campaign from Westinghouse to install GUV lamps in every American home^4^), it has remained mostly a niche technique in medical circles, in particular to reduce tuberculosis transmission.^5^ Research during the COVID-19 pandemic led to the conclusion that airborne transmission is dominant for this virus,^6^ and also important for other respiratory viruses.^7^ This has resulted in intense interest in methods to remove pathogens from the air, including ventilation, filtration, and air disinfection, in particular by GUV^.8^

GUV uses lamps that emit light in the UVC range (200-280 nm) to irradiate indoor air, which can inactivate aerosol-bound pathogens. It has traditionally been performed using filtered mercury lamps whose most intense emission is at 254 nm (“GUV254”). More recently the use of shorter wavelengths (“far UVC”, 200-230 nm) has gained in popularity, in particular using KrCl excimer with peak emission of 222 nm (“GUV222”). Extensive scientific reference information on GUV has been compiled at the online GUV Cheat Sheet.^9^

UVC lamps with wavelengths below 242 nm can generate O_3_,^10^ a dangerous pollutant. A recent review concluded that O_3_ generation by KrCl lamps was minimal: for example a 12 W lamp was estimated to take 267 h to produce 4.5 ppb O_3_ in a 30 m^3^ room in the absence of losses.^11^ A recent modeling paper estimated O_3_ generation to be nearly two orders-of-magnitude faster,^12^ but those findings have not been confirmed experimentally. There is also discussion in the literature whether O_3_ is mainly formed by the UVC radiation or by discharges in electrical connections^11^

In addition, it is typically difficult to quantify the GUV fluence rate that the air experiences in a room or chamber, since lamp emission results in inhomogeneous light spatial distributions, the reflectivity of materials at the GUV wavelengths varies widely, and due to continuous and highly variable air motion. Measurements in different points of a room to quantify the average, or computer modeling can be performed but are time consuming. Quantification of the radiation field with measurements of inactivation of viruses or bacteria require culture assays which are slow and very costly.

Here we present direct measurements of O_3_ production from KrCl excimer lamps in a laboratory chamber and compare them with literature estimates. A chemical tracer that allows quantification of GUV fluence rate is introduced. Measurements are also performed in an office. Significant O_3_ production is observed in both controlled-laboratory and real-world settings.

## 2. Methods

### Demonstration of tracers for GUV exposure of air

In this work, we use CBr_4_ as a chemical tracer of GUV exposure. We show that it has relatively fast decay under 222 nm irradiation and can be detected by commonly-available Proton-Transfer-Reaction Mass Spectrometers with high sensitivity. It does not react with common atmospheric oxidants such as O_3_, OH, or NO_3_ at typical indoor air concentrations. It has high vapor pressure and low water solubility which minimizes partitioning to room surfaces and tubing.^13,14^ More details can be found in Section S1.

### Laboratory chamber experiments

A well-characterized environmental chemical reaction chamber was used to measure the O_3_ production rate and CBr_4_ tracer decay for individual GUV fixtures. A ∼21 m^3^ Teflon reaction chamber (approximately 3×3×2 meters, LxWxH) is constructed of 50 μm thick FEP Teflon film (ATEC, Malibu, California). Temperature during the experiments was ∼20-25°C, and the built-in chamber UVA / visible lights were not used other than at occasional low-levels of visible lights for task lighting. The chamber systems are described in previous publications exploring chemical and physical processes of gases and aerosols.^15,16^ The GUV light source was placed either a few cm outside the chamber (at one corner shining into the bag and diagonally across to the opposite corner at mid-height) or placed within the chamber (at a lower corner mounted on a ring stand facing the opposite upper corner) (Fig. S1).

A typical experiment was conducted as follows. Prior to each experiment, the chamber was flushed for several hours with 400 LPM clean air (NO_x_<0.2 ppb; VOC < 50 ppb) from an AADCO generator (Model 737-15A) (at slightly positive pressure, 1-2 Pa) and then topped off to consistently reach the full volume (∼21 m^3^) by filling until the differential pressure reached 3.5 Pa. The GUV lamp was then turned on either continuously (Fig. 1b) or on/off in steps (e.g., 120 minutes on, 45 minutes off, Fig. 1a) for several hours. O_3_ formation was measured with a Thermo Scientific 49i O_3_ Analyzer. Later in the experiments while the GUV lamp was off, several ppb of CBr_4_ were added by placing the solid compound within a glass bulb and gently heating with a heat gun while flowing UHP nitrogen gas (for ∼2 minutes), and then mixing for 1 minute with a Teflon-coated mixing fan (integrated in the chamber). The on/off operation allowed to unequivocally attribute changes in the O_3_ and CBr_4_ mixing ratios to the GUV illumination, and to quantify any other losses separately. CBr_4_ was measured with Vocus (detected as CBr_3_^+^), which was calibrated by adding a known quantity to the chamber.^17^ See Sections S2 and S3 for more information on calibrations of the O_3_ analyzers and Vocus.

**Figure 1.**
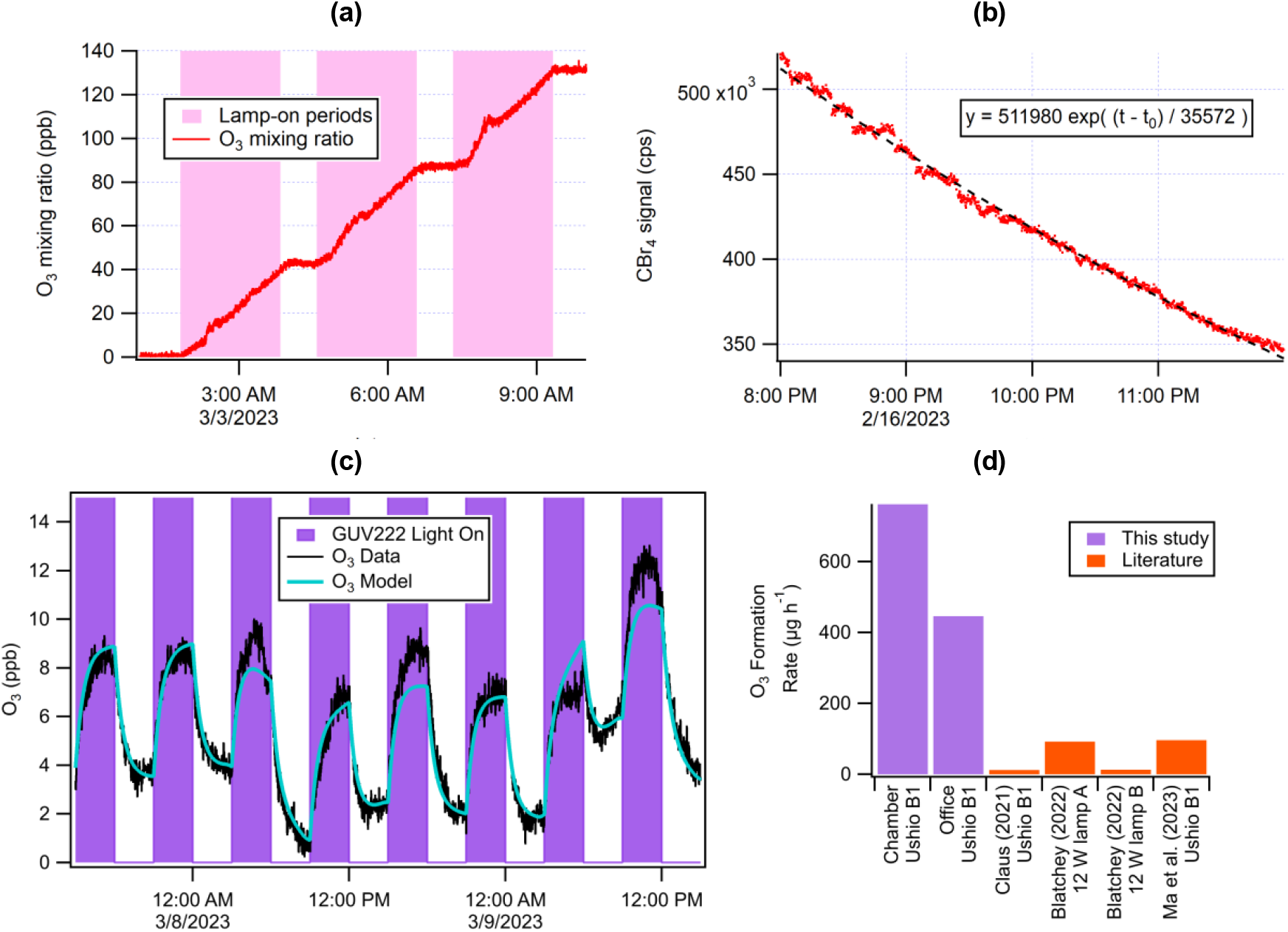
Time series of (a) O_3_ and (b) CBr_4_ during chamber experiments with the 12 W Far UV lamp (Ushio B1 module). (c) Time series of O_3_ in the office experiments, along with model results. (d) Comparison of O_3_ formation rates in this study with previous literature.

Since a single lamp fixture was illuminating from one corner of the bag, the fluence rate is not constant within the bag volume. However, on the timescales of the experiments (relative to the production/decay of the measured compounds), the air within the chamber is relatively well mixed. This is due to the continuous mixing that occurs in the absence of mechanical mixing, with a timescale of ∼10 minutes.^16^ This is apparent in the stepwise lamp illumination experiments, by the relatively quick stabilization of CBr_4_ and O_3_ when the light is turned off (Fig. 1a).

### Office experiments

Experiments were also performed in a small university office, which measured 4.0×2.7×3.1 m (LxWxH; vol. ∼33 m^3^). It has an entrance door and two windows. A supply and a return vent are located near the ceiling. To simulate a low-ventilation situation, the windows, gaps around utility penetrations, and supply / return vents were sealed with plastic sheeting or tape (Fig. S3).

The ventilation rate was quantified by monitoring the decay of CO_2_ after an injection (from a compressed gas cylinder) with an Aranet4 sensor (SAFTehnika, Latvia). A fan was turned on remotely for a few seconds after CO_2_ injection to ensure homogeneity within the room. The ventilation / infiltration rate was estimated with an exponential fit to the CO_2_ decay to be as low as ∼0.44 h^-1^ (timescale of ∼2.3 h, comparable to typical residences),^18^ but with some experiment-to-experiment variability. For this reason CO_2_ was injected also during the O_3_ decay or production experiments.

To quantify the O_3_ decay to surfaces and to gas and aerosol chemistry in the room, the O_3_ decay in the room was measured with a 2B 205 analyzer. The decay was fit to an exponential, and the O_3_ deposition rate coefficient was determined by subtracting the ventilation rate coefficient (Fig. S4).

## 3. Results

### Theoretical estimation of O_3_ production and tracer decay

In this study, we tested lamps from different manufacturers (Table 1). The emission spectrum of the Ushio B1 lamp that is used by multiple lamp manufacturers is available from NIST (Fig. S2). The absorption spectra of O_2_ and CBr_4_ are well-known.^10,19^ Their expected photolysis rates under the Ushio B1 lamp irradiation can be calculated by integrating the product of UV fluence rate and absorption cross sections of O_2_ or CBr_4_ over the wavelengths of interest. As 2 molecules of O_3_ are produced per O_2_ molecule photolyzed, the theoretical O_3_ production rate for the Ushio B1 lamp at a total UV fluence rate of 2.3×10^12^ photons cm^-2^ s^-1^ is ∼22 ppb h^-1^. At the same UV intensity, the theoretical CBr_4_ photolysis rate is 0.097 h^-1^. The ratio between them (P_O3_/J_CBr4_ ∼ 230 ppb), i.e. O_3_ production through O_2_ photolysis over a period for an e-fold decay of CBr_4_, is independent of UV fluence rate and is characteristic of a specific GUV222 lamp.

**Table 1.**
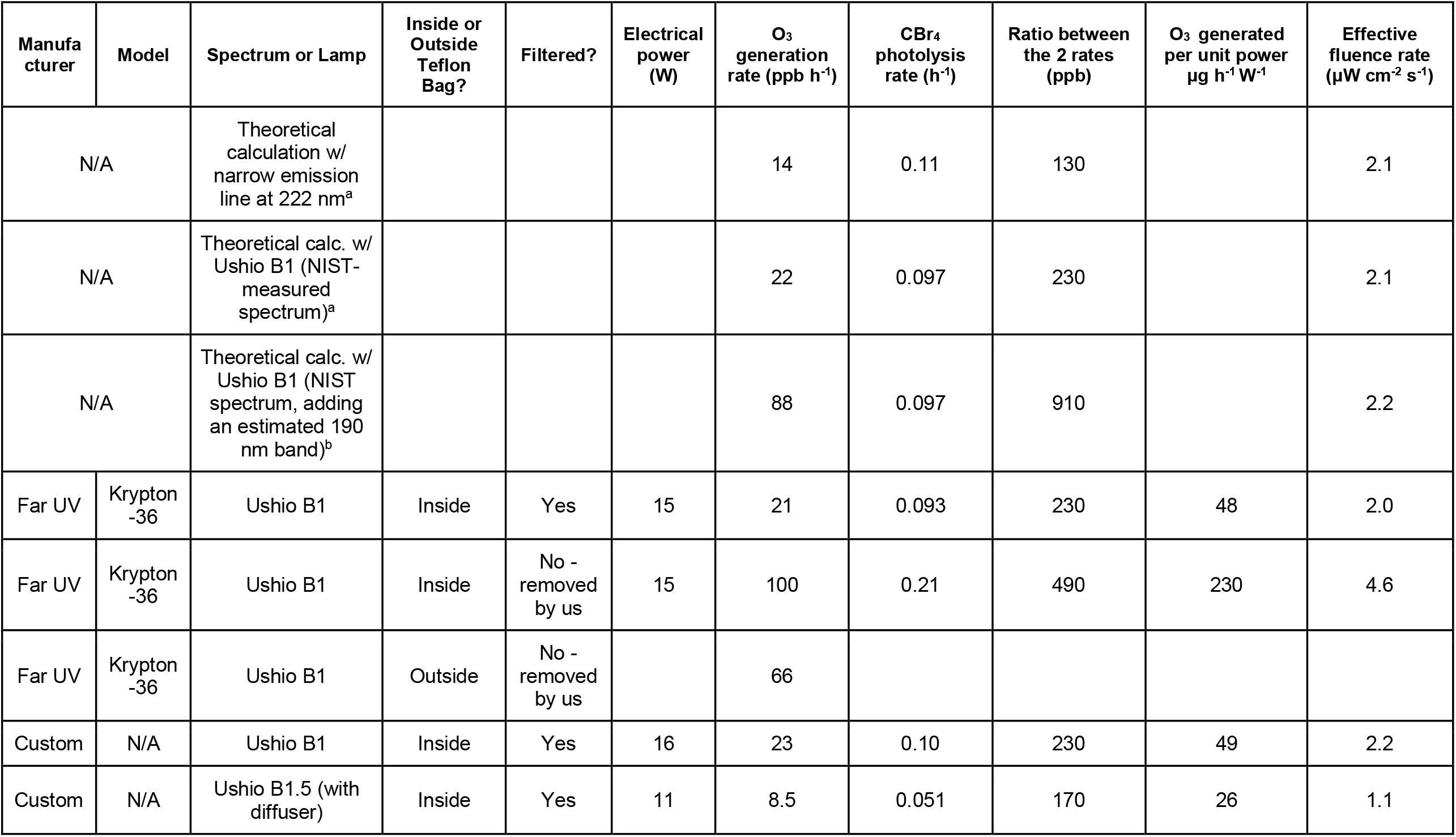
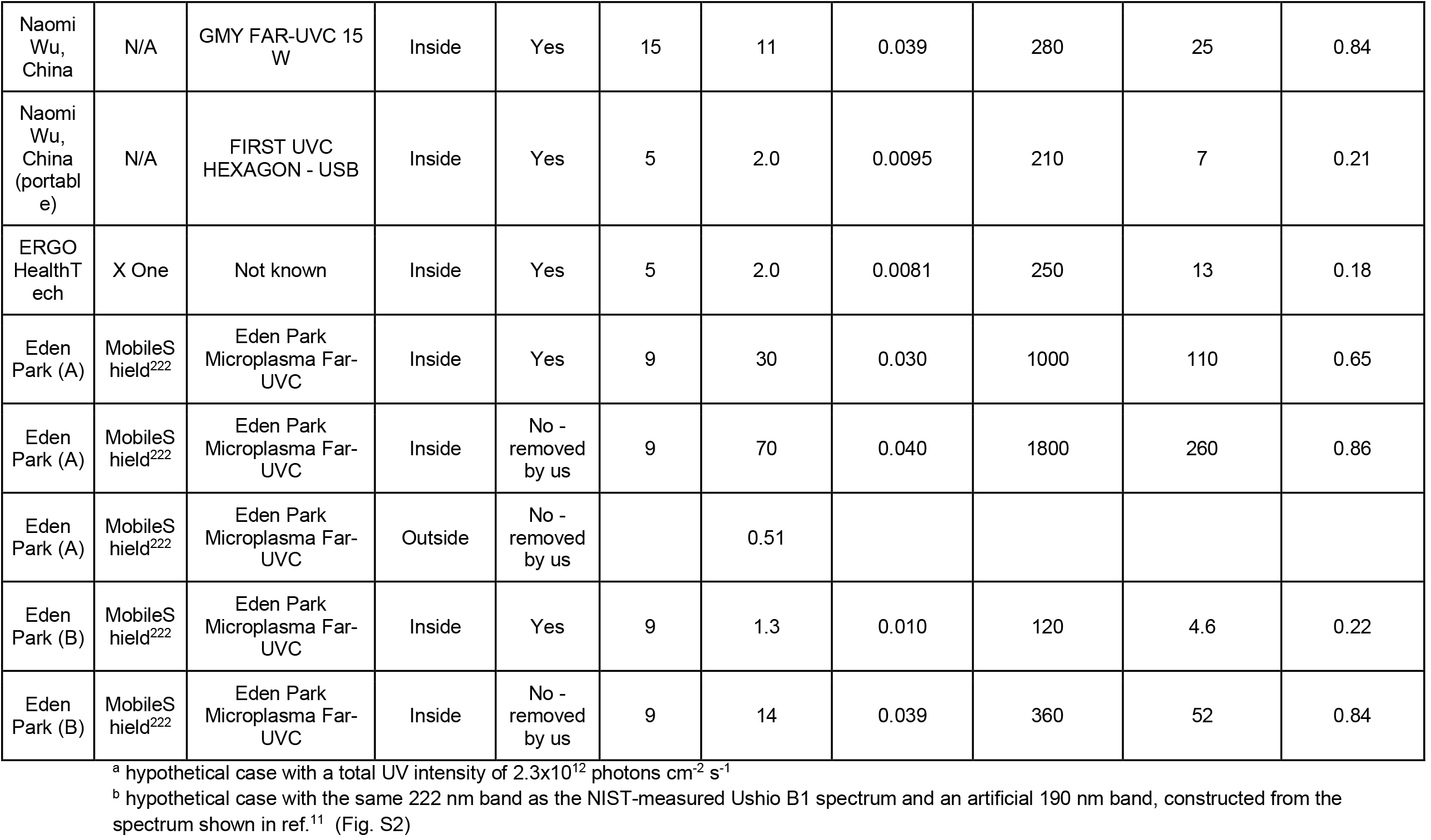
KrCl excimer lamps tested in the chamber experiments and key results. Also shown are the results of first-principles calculations with different lamp spectra.

When unfiltered optically, the emission of KrCl excimer lamp also includes a band centered at 190 nm (Fig. S2).^11^ If this band is added to the theoretical calculation (as a proxy of unfiltered lamps), the O_3_ generation rate is increased by a factor of ∼4. Although the 190 nm band has much lower intensity, the absorption cross section of O_2_ is on average ∼2 orders of magnitude larger for the 190 nm band than for the 222 nm one. However, this band has little impact on CBr_4_ as its cross section below 200 nm is much lower. This results in a higher P_O3_/J_CBr_ ratio (∼900 ppb) than for the filtered lamp spectrum.

### O_3_ production and CBr_4_ in the chamber

Results of a typical chamber experiment (Ushio B1) are shown in Fig. 1a. O_3_ increases approximately linearly with time when the lamp is on. When this lamp is on for an extended period (days), O_3_ in the chamber can reach ppm levels (Fig. S5). At very high O_3_ concentrations, the small loss rate coefficient of O_3_ (mainly due to O_3_ photolysis by the 222 nm band, Fig. S6) slightly reduces the rate of O_3_ increase. An Ushio B1 lamp generates ∼22 ppb O_3_ per h, very close to the theoretical case shown above. The effective UVC fluence rate inferred from the CBr_4_ photolytic decay rate (∼0.1 h^-1^, dilution corrected, Table 1) (Fig. 1b), is also very close to the theoretical case value. P_O3_/J_CBr4_, a characteristic of the lamp, is almost the same as the theoretical case value (Table 1).

The other devices tested in this study, with electrical power ranging from ∼5 W (portable device) to ∼15 W, also have P_O3_/J_CBr4_ values in the range of 200-300 ppb, indicating similar spectral characteristics of their emissions. The exceptions are the lamps whose filters were removed for our tests, two Eden Park lamps we tested, and an Ushio B1.5 lamp with a diffuser.

The Far UV device with filter removed has 4 times more O_3_ production and >100% larger CBr_4_ decay than when it has the filter, leading to about twice P_O3_/J_CBr4_ of the device with the filter. Without the filter, more photons of the 222 nm band are allowed out of the device, leading to faster CBr_4_ decay. The 190 nm band is also unfiltered, producing much more O_3_ than the 222 nm band of the device without filter can produce.

The Eden Park lamp has almost the same emission spectrum as the Ushio B1 (Fig. S7). With its filter, the first Eden Park device tested (EP-A, Table 1) results in ∼1/3 CBr_4_ decay vs. Ushio B1, while producing more O_3_. Most of this unexpectedly high O_3_ production appears to be due to electrical discharge within the electrical components of the device (but outside the lamp). We arrive at this conclusion after additional tests: (i) low O_3_ production by the EP-A in the chamber bag when located outside the bag, in contrast to the Ushio B1 module (Table 1). (ii) For O_3_ measurement just outside the EP-A housing, but not in front of the light, the O_3_ monitor detects ppm-level O_3_ (Fig. S8), implying very strong non-photochemical O_3_ production inside the device.

In contrast, the other Eden Park device test (EP-B) did not produce an excessive amount of O_3_ in its housing, implying no undesired electrical discharge there. It also only produces 1.3 and 14 ppb O_3_ per h in the chamber with and without its filter, respectively, resulting in significantly lower P_O3_/J_CBr4_ than the Ushio B1 lamps. The reasons for the lower P_O3_/J_CBr4_ of the Ushio B1.5 module and EP-B lamp are unclear, as they have similar emission spectra to Ushio B1 (Fig. S7).

### O_3_ mass balance in an office

The Far UV lamp was repeatedly cycled on (3 h) and off (3 h) together with periodic CO_2_ injections (Fig. 1c and Fig. S9). O_3_ rapidly rose once the lamp was turned on and reached an approximate steady state (8-14 ppb, typically increasing ∼6.5 ppb). After turning off the lamp, O_3_ rapidly decreased, also quickly reaching a steady state. Background O_3_ in the office, as indicated by the steady-state O_3_ level at the end of lamp-off periods, varied by ∼4 ppb during the experiment. It is affected by ventilation rate, deposition, as well as O_3_ in outdoor/adjacent-room air infiltrating into the office. Ventilation rates ranged 0.62-0.96 h^-1^ (Fig. S9). O_3_ deposition rates were more variable (range 0.5-2.3 h^-1^, average 0.78 h^-1^, Fig. S9).

O_3_ in the office was modeled with a chemical-kinetics simulator.^20^ The model was constrained by the measured O_3_ and CO_2_ concentrations and decays (Section S4). The measured and modeled O_3_ are in good agreement (Fig. 1c). The O_3_ production rate of the Ushio B1 module in the office (Fig. S3) is estimated to be 8.6 ppb/h from the constrained model. This is ∼39% of the value measured in the chamber, which is explained by the larger volume of the office (∼32.9 vs. ∼20.6 m^3^) and the shorter effective UV pathlength (∼3.2 vs. ∼4.5 m). Scaling results in a difference of 12%, thus showing agreement well within experimental uncertainties (Fig. S10).

### Implications

Significant amounts of O_3_ can be produced by GUV-222 lamps in both controlled-laboratory and real-world settings. Our results of 762 μg h^-1^ for a 21 m^3^ chamber and 446 μg h^-1^ for an office with a shorter light path are summarized in Fig. 1d. Note that these results would be ∼18% higher at sea level due to the reduced ambient pressure in Boulder. For comparison, previous reports for the same GUV222 module (or modules using the same electrical power) from the literature of 12,^11^ 13 and 92,^21^ and 96 μg h^-1 22^ are also shown. These had been used to conclude that O_3_ generation from GUV222 is not a concern. On average, our results are an order of magnitude larger than the literature. The discrepancy may arise because most prior measurements were performed in small boxes, with very short optical pathlengths and high surface/volume ratios that are not representative of real room applications. The former may lead to smaller O_3_ production rate, and the latter to substantial losses to the box surfaces, which were not accounted for. Moreover, some of these measurements may have been made with low-cost electrochemical O_3_ sensors. We tested four sensors and found them to be unresponsive to O_3_ mixing ratios below 200 ppb, therefore such sensors are not useful for this problem (Section S5).

O_3_ itself is a major air pollutant with excess deaths observed at levels below those in occupational guidelines of 50-100 ppb.^23,24^ Critically, it can also lead to formation of other pollutants including particulate matter,^12^ which has ∼10-30 times higher excess death impacts on a mass basis (Section S6).^23,25^ O_3_ production by GUV222 lamps thus can be a major concern, at least under low-ventilation conditions.

Our experiments have an average fluence rate of ∼2.1 μW cm^-2^ s^-1^, about ⅓ of the recently-updated ACGIH eye limit, and approximately consistent with a room installation that achieves the ACGIH limit at eye level (H. Claus, pers. comm.). ACGIH should consider reduced limits at low ventilation rates. Current literature estimates of the GUV222 disinfection rate coefficient for SARS-CoV-2 span a factor of 33.^26,27^ Future research should focus on narrowing down this range, which may allow high efficacy of GUV222 at lower fluences, thus reducing impacts on indoor air.

## Data Availability

All data produced in the present work are contained in the manuscript

## Acknowledgements

We thank the Balvi Filantropic Fund, the CIRES Innovative Research Program, and the Sloan Foundation (grant 2019-12444) for support of this work. We thank Shelly Miller, Kenneth Wood, Ewan Eadie, Catherine Noakes, Dustin Poppendieck, Michael Link, Mikael Ehn, Jesse Kroll, Victoria Barber, John Balmes, and the larger GUV scientific and technical communities for useful discussions. We are grateful to Holger Claus, Aaron Collins, Matthew Pang, Kristina Chang, Scott Diddams and Naomi Wu for sharing data or materials and useful discussions.

## Supporting Information

### Supporting Information Text Sections

#### Section S1. Selection and Application of CBr_4_ a tracer of GUV fluence rate in air

A useful chemical tracer of GUV exposure should not react (or react slowly) with common atmospheric oxidants such as O_3_, OH, or the NO_3_ radical at typical indoor air concentrations. O_3_ and NO_3_ typically react only with C=C double bonds, while OH can abstract hydrogens from most organic molecules.^1^ A tracer should also have a high absorption cross section at the most common GUV wavelengths (222 and 254 nm), so that its decay is large enough and can be quantified over reasonable time scales despite instrumental noise. It should have high vapor pressure and low water solubility to reduce partitioning to room surfaces and tubing.^2,3^ It should not be highly toxic, and it should be detectable with high sensitivity with existing instrumentation, so that its mixing ratio can be kept low to minimize any unwanted effects on chemistry or human exposure concerns. After comparing a few candidate species, we selected CBr_4_ as a tracer. We show that it has relatively fast decay under 222 nm irradiation and can be detected by a commonly-available Proton-Transfer-Reaction Mass Spectrometer with high sensitivity.

A search for species with these properties that can serve as a GUV fluence rate tracer at both main GUV wavelengths in use (222 and 254 nm) identified three candidates, shown in the table below. Other species considered (including CF_2_Br_2_, CCl_3_Br, CF_2_I_2_, C_2_F_5_I, CF_3_I, OCS, and diacetyl) had too low absorption cross section (*σ*) at one of the key GUV wavelengths. CBr_4_ was selected due to having the highest *σ* (and thus the fastest photolysis rates), low reactivity with oxidants, and being detectable with the Vocus instrument with high sensitivity. This instrument is widely-available in air chemistry research laboratories. This molecule is an excellent tracer in particular for GUV222, as its absorption cross section is highest at that wavelength, and falls about an order of magnitude when 10 nm away on either side of the peak. The absorption cross sections of CBr_4_, O_2_ and O_3_ are shown in Fig. S6.

To quantify the sensitivity of the Vocus to CBr_4_, 20.10 mg CBr_4_ was evaporated under clean nitrogen flow into the chamber (whose volume was measured by quantitatively injecting CO_2_ and measuring the concentration). A teflon-coated fan was run for one minute following the addition to ensure complete mixing. The concentration of CBr_4_ in the chamber and measured ion counts per second (cps) for the CBr_3_^+^ ion were used to determine the sensitivity in cps ppb^-1^.

As CBr_4_ also absorbs at 254 nm, it can cause interferences in the Thermo Scientific 49i O_3_ Analyzer, which uses absorption at 254 nm to measure O_3_. We measured the apparent O_3_ signal in the Thermo Scientific 49i O_3_ Analyzer at several CBr_4_ concentrations in the absence of O_3_ in the chamber. Below 200 ppb CBr_4_, the interference of CBr_4_ is approximately linear with its concentration (Fig. S11). The O_3_ signal due to CBr_4_ interference is ∼0.007 ppb per ppb CBr_4_ in this CBr_4_ concentration range, in which most of the experiments in this study were (usually on the range 1-10 ppb). At very high CBr_4_ concentration (∼500 ppb), the relationship between the concentration and the O_3_ interference is no longer linear.

During the O_3_ generation rate quantification experiments, CBr_4_ (Sigma-Aldrich) was added to the chamber after the O_3_ quantification was done. This order was followed because CBr_4_ photolysis produces Br radicals that can catalytically destroy O_3_ in a similar way as catalytic destruction of stratospheric O_3_ by Cl.^4^ In the presence of CBr_4_ and GUV irradiation (and hence Br atoms), a steady state for O_3_ exists that is governed by Br concentration (and hence CBr_4_ and GUV fluence rate). Fig. S12 shows the O_3_-CBr_4_ relationship during a long CBr_4_ decay experiment with the Far UV fixture (with filter). CBr_4_ decay was relatively slow. Therefore, O_3_ concentration responded to CBr_4_ relatively rapidly and could be regarded as steady-state concentration.

**Table S1.** key properties of potential GUV average fluence rate tracers. (*): Lifetimes are estimated for typical indoor GUV intensities of 2.61 × 10^12^ and 1.06 × 10^14^ photons cm^-2^ s^-1^ at 222 and 254 nm, respectively, and for an OH concentration of 1.5 × 10^6^ molec. cm^-3^. (**): no specific exposure limit, hazard information available at https://pubchem.ncbi.nlm.nih.gov/.

| Species | CBr<sub>4</sub> | CHBr<sub>3</sub> | BrCOC<br>OBr | CF<sub>2</sub>Br<sub>2</sub> | CCl<sub>3</sub>Br | CF<sub>2</sub>I<sub>2</sub> | C<sub>2</sub>F<sub>5</sub>I | CF<sub>3</sub>I |
| --- | --- | --- | --- | --- | --- | --- | --- | --- |
| σ at 222 nm | 2.85E-18 | 1.36E-18 | 2.00E-18 | 3.68E-19 | 4.60E-19 | 1.12E-18 | 4.73E-19 | 4.26E-19 |
| σ at 254 nm | 1.32E-17 | 5.78E-18 | 7.00E-18 | 2.44E-18 | 4.80E-19 | 6.00E-19 | 2.48E-20 | 2.06E-20 |
| OH rate coeff. | - | 2.70E-13 | - | - | - | - | - | - |
| GUV-222<br>lifetime* (h) | 8.0 | 18.4 | 15.2 | 43.5 | 221.4 | 177.0 | 4284 | 5158 |
| GUV-254<br>lifetime* (h) | 0.9 | 1.9 | 1.3 | 7.1 | 5.7 | 2.3 | 5.6 | 6.2 |
| OH lifetime* (h) | - | 686 | - | - | - | - | - | - |
| Exposure limit<br>(ppb) | 100 | 500 | ** | 10<sup>5</sup> | ** | ** | ** | ** |

#### Section S2. Selection of acetone as a tracer of Vocus sensitivity

A Vocus sensitivity tracer was useful for some experiments with weak lamps, e.g. the Naomi Wu (portable) and Eden Park (B) devices (Table 1), where the CBr_4_ photolytic decay was small (∼0.01 h^-1^) and the Vocus sensitivity drift could be of a comparable magnitude.

Acetone was selected as a tracer of Vocus sensitivity because of the following properties. First, the Vocus instrument detects acetone with high sensitivity.^5^ Besides, its absorption cross section drops by orders of magnitude between 195-200 nm and is 3-4 orders of magnitude lower than that of CBr_4_ (Fig. S6), leading to little photolysis by the GUV band centered at 222 nm.

Moreover, it is unreactive with O_3_, and its reaction with OH is negligible under the conditions in this study. After injection into the chamber, the acetone signal can serve to continuously quantify small variations in Vocus sensitivity, for the experiments where the GUV device has the optical filter that filters the 190 nm band.

#### Section S3. Calibration of O_3_ analyzers used for chamber and office experiments

O_3_ formation in the chamber was always measured with a Thermo Scientific 49i O_3_ Analyzer. That analyzer was calibrated using actinometry within the experimental chamber, where ∼40 ppb of NO_2_ was injected into the dry chamber, and the UVA lights are stepped through four discrete levels (between 10-100% of total UVA power). Equal amounts of NO and O_3_ are generated, which are monitored with the O_3_ analyzer and a Thermo Scientific 42i-TL NO-NO_2_-NO_x_ Analyzer. The NO_x_ analyzer was calibrated using a NIST-certified (±2%) calibration standard (gas cylinder with NO in N_2_) and Thermo Scientific Multi-Gas Calibrator (146i). We estimate that this method provides a calibration accuracy of ±5% for the O_3_ analyzer.

O_3_ decay rates and concentrations in the office experiments were always measured with a 2B Model 205 analyzer, which was cross-calibrated with the Thermo analyzer used in the chamber experiments, with its zero calibrated with zero air (resulting accuracy of ±7%, and zero uncertainty of ± 0.5 ppb).

#### Section S4. Data analysis and kinetic modeling for the office O_3_ production experiment

##### Characterization Tests

In characterization tests (without a GUV lamp), the ventilation rate was measured as 0.52-0.61 hr^-1^ (τ: 1.6-1.9 h) using CO_2_ decay. For initial characterization of O_3_ decay, ∼400-500 ppb O_3_ were generated with an unfiltered low-pressure Hg lamp with partial emission at 185 nm (BHK 82-9304-03) (Fig. S3), together with CO_2_ injection. As expected, the decay of O_3_ generated by the Hg lamp was faster than CO_2_ decay (Fig. S4), because of other O_3_ losses than ventilation (dry deposition, reactions with VOC emitted indoors etc.). Subtracting the two rate coefficients yields an O_3_ deposition coefficient of 0.76-1.1 h^-1^ (τ: 0.93-1.3 h), and the overall O_3_ decay rate as 1.3-1.7 hr^-1^ (τ: 0.59-0.78 h).

Given the variability in these experiments, the CO_2_ and O_3_ decay rates were measured after each period in which the GUV lamp was turned on, as described in the main paper.

##### Modeling

To quantify the O_3_ production rate from the GUV lamp, all relevant parameters affecting O_3_ concentration in the office were modeled in KinSim. The first-order ventilation rate, first order deposition rate coefficient (implicitly including gas and aerosol reactions), and the approximate mixing ratio of O_3_ entering the room from outside the room had to be measured or estimated.

The ventilation rate was directly measured using CO_2_ pulse injection experiments discussed in the main paper, and the deposition rate was estimated by subtracting the ventilation rate from the first-order overall O_3_ loss rate coefficient (green fit lines in Fig. S9). From here, the effective value for the O_3_ mixing in from outside the room was approximated through tuning of the model when the lamp was off (blue points in Fig. S9). Finally, across 5 of the 8 peaks the production rate was tuned individually until it matched with each peak, and then the average was used as a constant production rate (individual values shown in Fig. S9 and used as a metric of uncertainty). Peaks 3, 7, and 8 were excluded due to rapidly changing O_3_ background levels. As it was found that the estimated O_3_ deposition rate varied substantially for the different light cycles, it was assumed to be constant for the model and the average value was used as an input (and computing “outside” O_3_). This choice was made since, given the relative invariability of the ventilation rate and lack of activity in the room, it seemed unlikely that actual O_3_ deposition rate coefficient would change all that much. More likely, the variability was more driven by a combination of uncertainty in changing ventilation rates and “outside” O_3_ on timescales faster than these parameters could be quantified, as well as the uncertainties associated with fitting and subtracting decay and ventilation rates. Figure S9 displays all of these parameters in the first “variable deposition” scenario as well as the second “constant deposition scenario”. The O_3_ production rate for the GUV lamp was only calculated for the second scenario.

As seen in Fig. S10, The O_3_ production rate for the conference room is slightly lower than that of the chamber due to the size of the room (∼32.9 vs. ∼20.6 m^3^) and effective UV pathlength (∼3.2 vs. ∼4.5 m). However, the results are within error bars when these differences are taken into account. The effective path length is shorter in the conference room both because of the shorter length of the room (3.8 m), and the combination with the narrowness of the room and furniture obstructions, which we estimate to reduce effective pathlength by ∼15%.

#### Section S5. Evaluation of handheld electrochemical O_3_ monitors

Three low-cost (∼$100) handheld electrochemical O_3_ monitor models were compared with our research-grade UV absorption Thermo Scientific Model 49i Ozone Analyzer. Table S2 lists all three monitors with their relevant information and specs. Two identical monitors were tested for the Shenzhen Dienmern model.

**Table S2.**
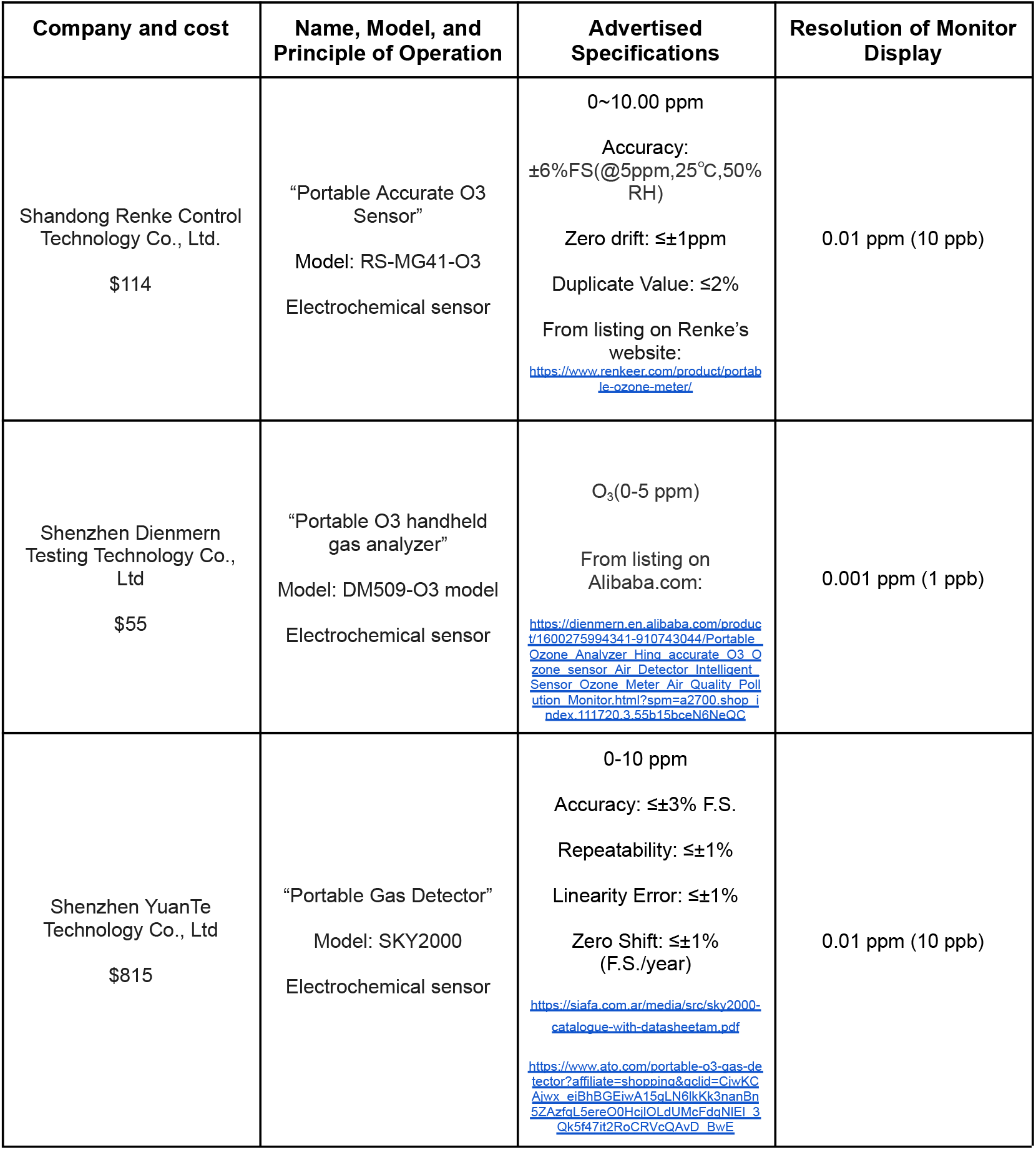
Specifications for all low-cost O_3_ monitors tested in the chamber instrument.

The set up for these monitors in the chamber can be seen in Fig. S13. O_3_ was injected into the chamber with a commercial O_3_ generator (BMT 802N) periodically, followed by mixing with a fan, in order to generate constant concentration O_3_ “steps”. The Thermo O_3_ concentrations were logged continuously and the concentrations of the hand-held O_3_ monitors were manually read and recorded for each step. The results of this comparison can be seen in Figs. S14 and S15. Performance is very poor at the relevant levels for typical indoor O_3_ and the levels expected when GUV222 is applied at e.g. ACGIH limits (i.e. no response for O_3_ <150–400 ppb). At higher levels > 200 ppb, the Shenzhen YuanTe monitor eventually quantifies O_3_ with good accuracy, while the other two models continue to be low by a factor of ∼8. The Shenzhen YuanTe monitor is also distinct from the other two as it is a factor of ∼10 more expensive. According to the information that we could find (see Table S2), two of these monitors appear to have failed their accuracy and/or zero drift specifications. For the other one, the only specs that we found were the measurement range and statements that it has high accuracy. To the best of our current knowledge, the lowest cost monitors capable of accurate O_3_ measurements at single-digit or tens of ppb-level concentrations are based on UV absorption, and cost at least $6000.

#### Section S6. Comparison of the health effects (premature death) for O_3_ and fine PM

The mortality due to long-term exposure to O_3_ (per unit mass of O_3_) can be estimated from the literature. Turner et al. (2016)^6^ is considered the best study to date on this topic (J. Balmes, UCSF, pers. comm., 2023). This study reports an increase in all-cause mortality of 2% per 10 ppb increase in O_3_. 10 ppb are equivalent to 19.7 μg m^-3^ at 1 atm and 298 K. Thus, we can estimate the risk per unit mass of O_3_ as 2% / 19.7 = 0.10% per μg m^-3^.

For comparison, the mortality due to long-term exposure to PM_2.5_ can be estimated from Figure 2a of Weichenthal et al. (2022)^7^. For their updated exposure function, the increased relative risk of mortality per unit increase in PM_2.5_ (i.e. the slope of the curve) is highest between 2.5-4 μg m^-3^, at about 3.2% per μg m^-3^. At concentrations around the US PM_2.5_ average of ∼7 μg m^-3^, this value is 1% per μg m^-3^ for their updated function, and 0.95% per μg m^-3^ for the prior literature function.

Thus depending on the estimate used for PM_2.5_ risk, the all-cause mortality risk of PM_2.5_ is 9.5-32 times larger than for O_3_.

## Supporting Information Figures

**Figure S1.**
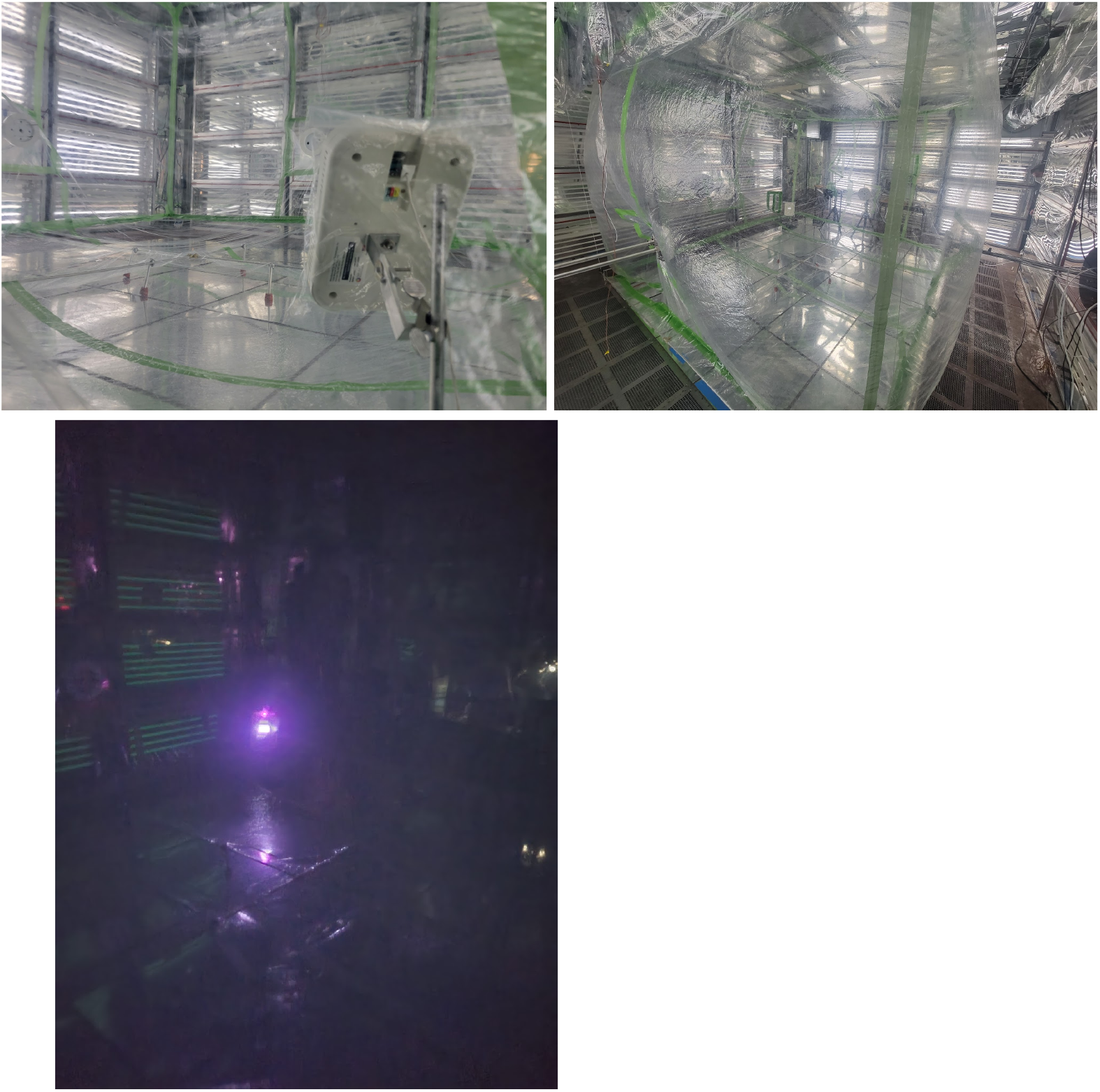
Pictures showing the FarUV GUV222 lamp mounted inside the Teflon chamber. Other lamps were tested in the same physical configuration. All tests were performed with the visible lights off, as in the last picture.

**Figure S2.**
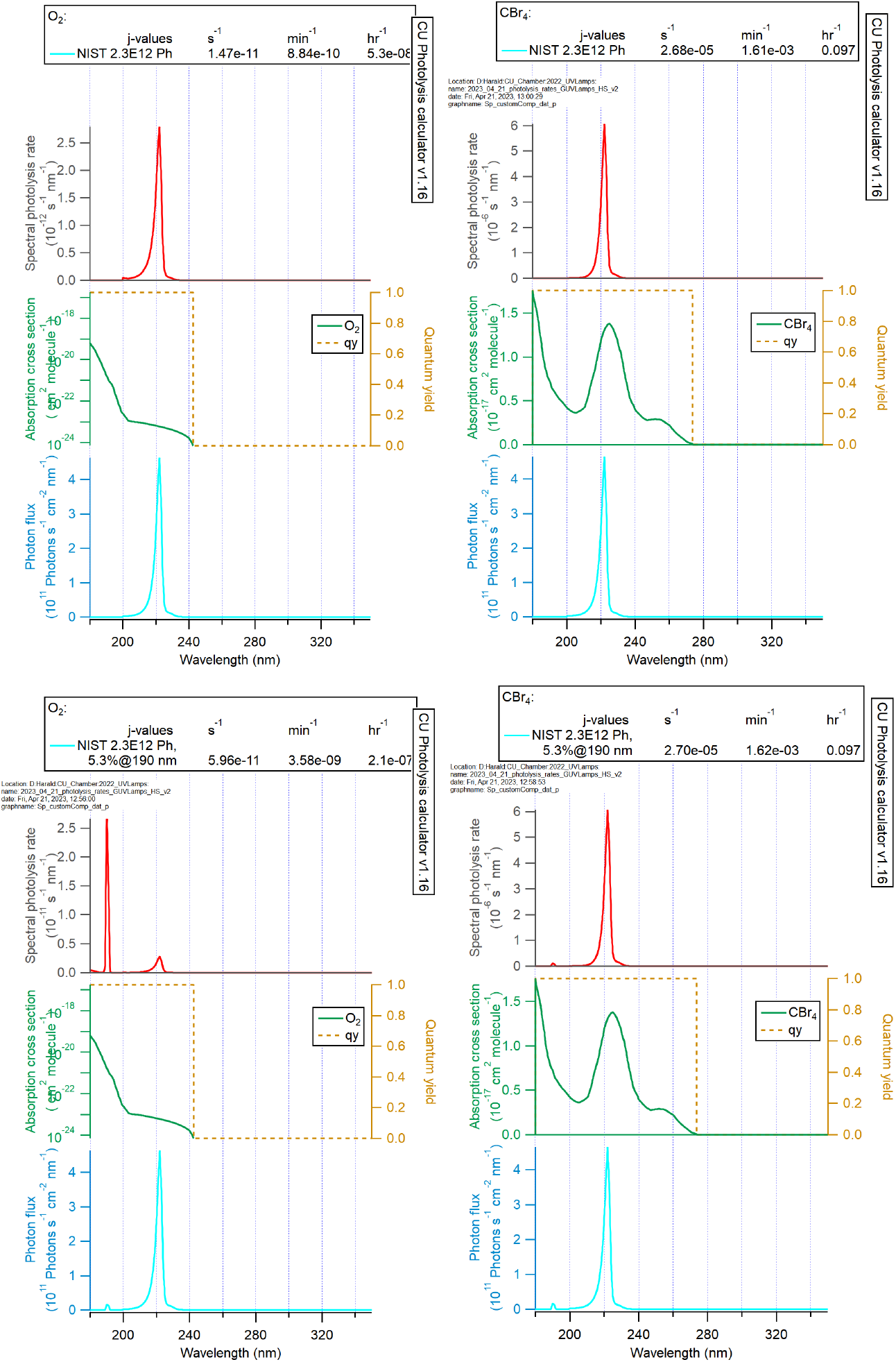
(Top) photolysis spectra and rates from O_2_ (left) and CBr_4_ (right) for NIST-measured Ushio lamp spectrum. (Bottom) results for the same lamp with an additional peak of 5.3% of the peak intensity manually added centered at 190 nm, estimated from Claus (2021).^8^ These results were generated with the CU-Boulder photolysis calculator.

**Figure S3.**
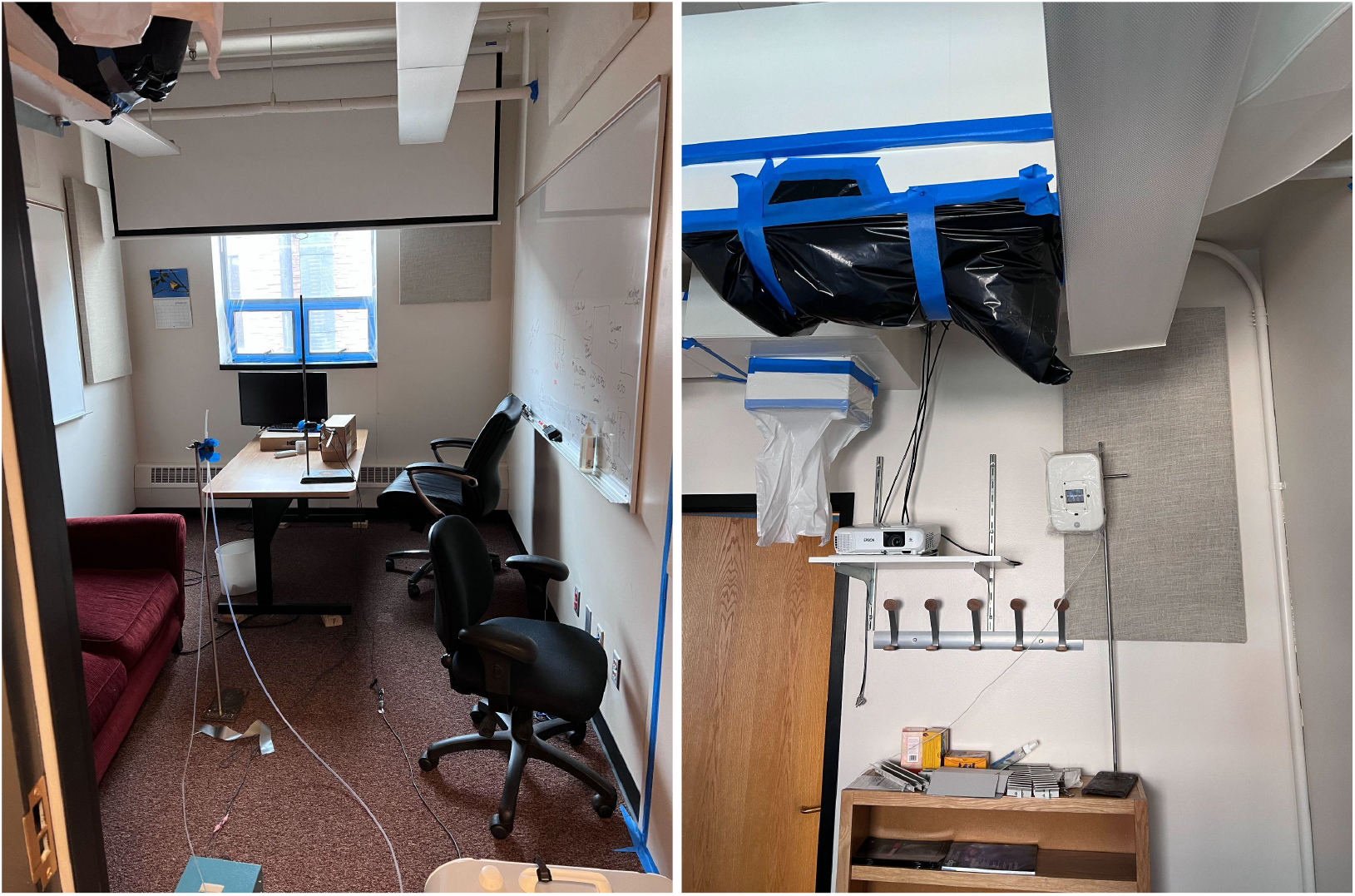
Experimental setup in the test office. The O_3_ sampling tube and CO_2_ injection tube were placed in the middle of the room on a ring stand (left). The GUV lamp was placed high in the room against the West wall of the room (right). The path of the light was interrupted by the furniture and walls, and the effective pathlength in the main paper was estimated to account for those obstructions.

**Figure S4.**
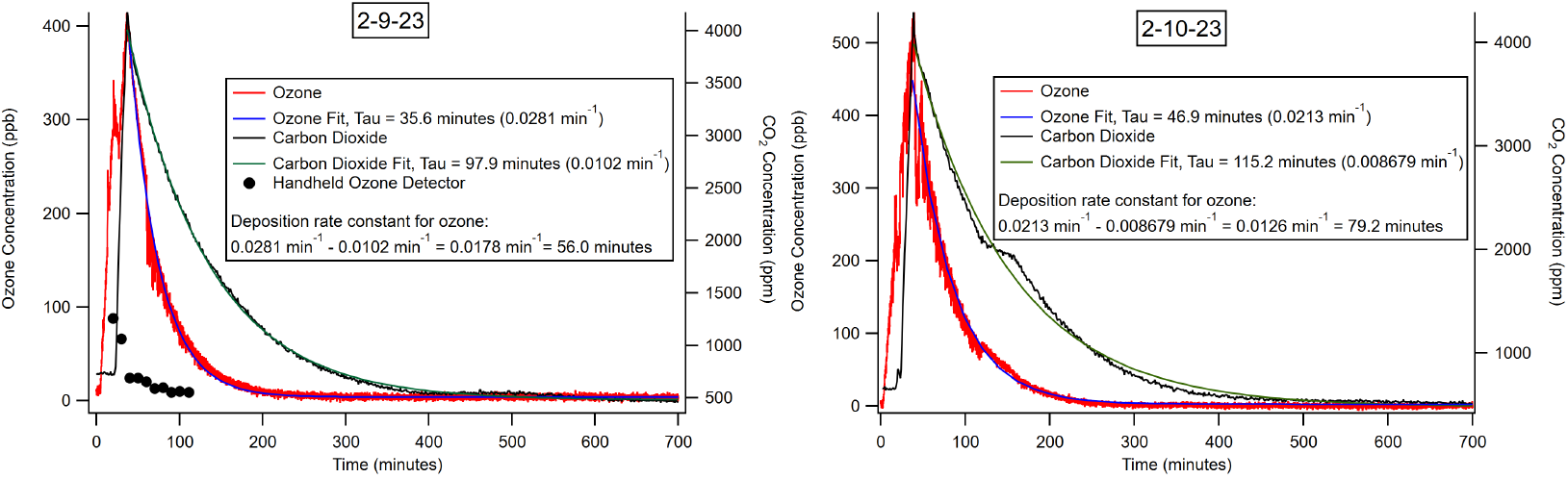
Decays of O_3_ and CO_2_ in the office experiments, along with the approximate decay first-order rate coefficients for 2 experiments on 2 different days. Measurements from a handheld low-cost O_3_ detector are also shown, which underestimated the O_3_ concentration by about an order-of-magnitude (see Section S5).

**Figure S5.**
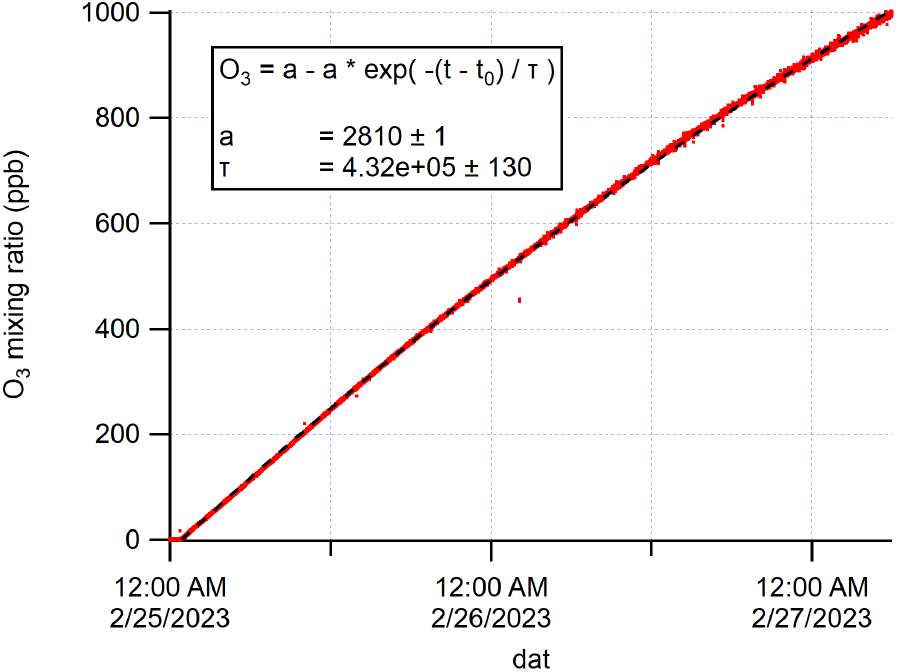
O_3_ concentration vs. time in the chamber when the custom lamp with an Ushio B1 module was turned on for an extended period.

**Figure S6.**
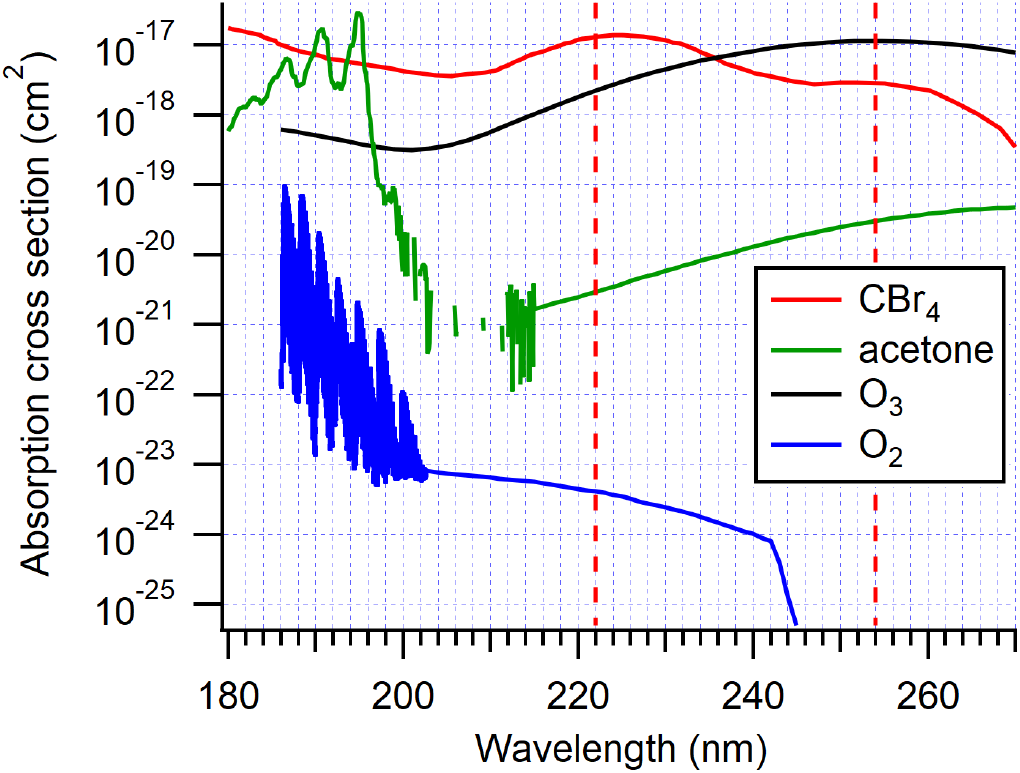
Absorption cross sections vs. UV wavelength for O_2_, O_3_, acetone and CBr_4_.^9,10^

**Figure S7.**
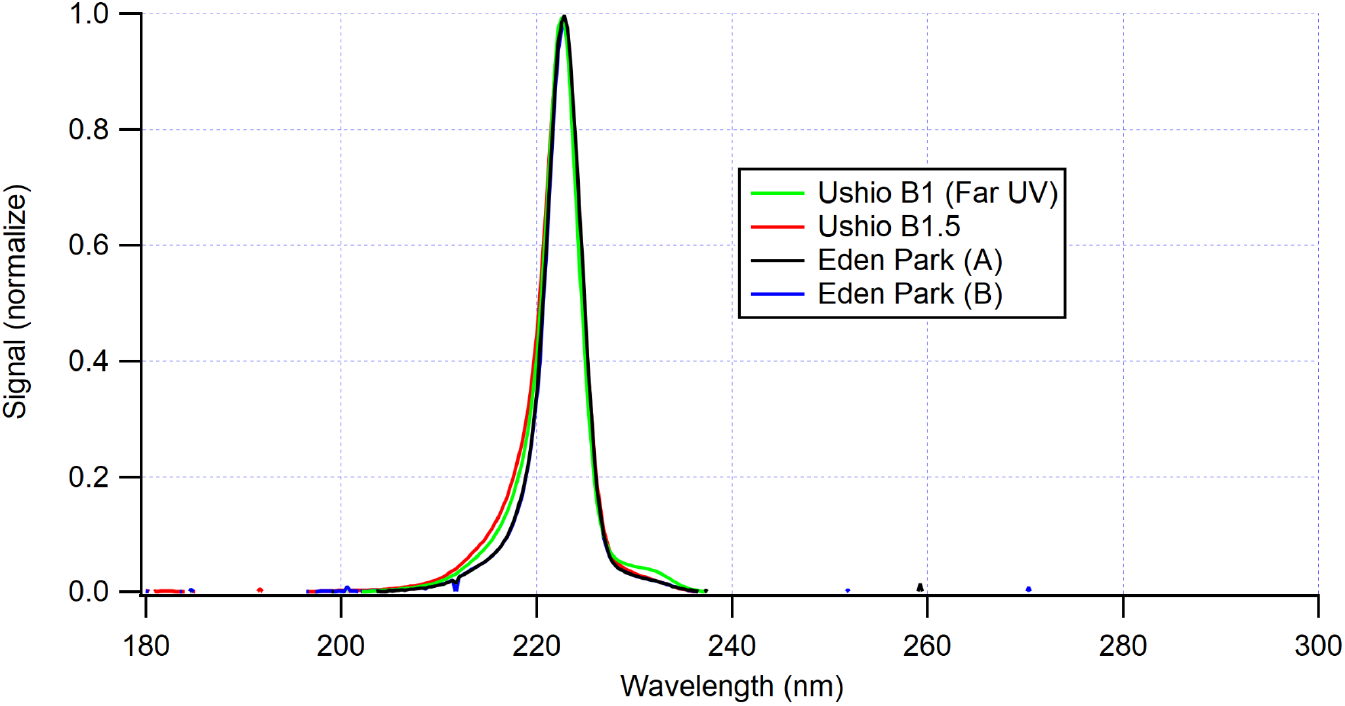
Measured emission spectra of the Far UV (Ushio B1), Ushio B1.5, and Eden Park (A) and (B) lamps. All spectra were measured with their original filters in place.

**Figure S8.**
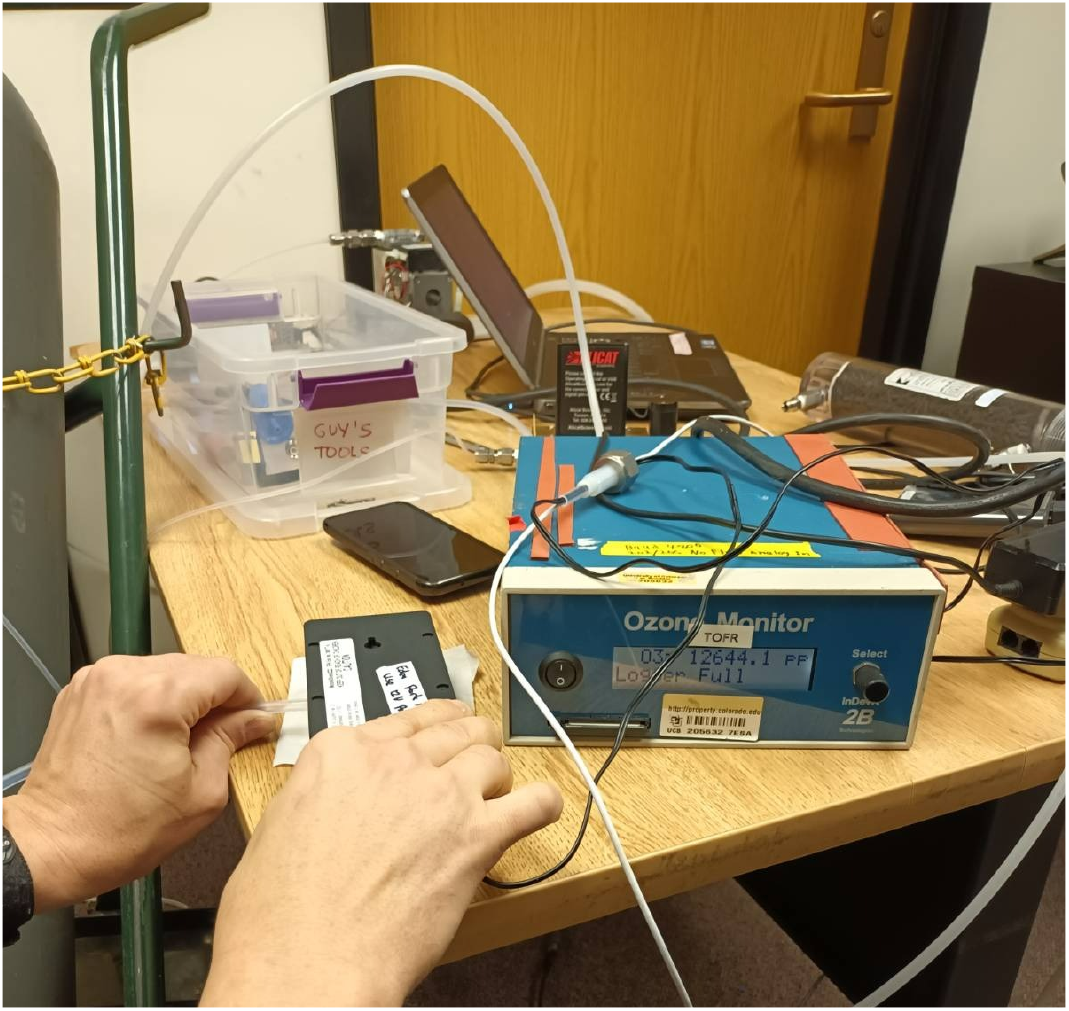
Picture of the setup for O_3_ measurement just outside the electronics compartment of the Eden Park (A) device (black box held with right hand). The light emission surface points down into the table. The 2B O_3_ analyzer displays a measured O_3_ concentration of 12.6 ppm. Similar readings were observed for a period of several minutes.

**Figure S9.**
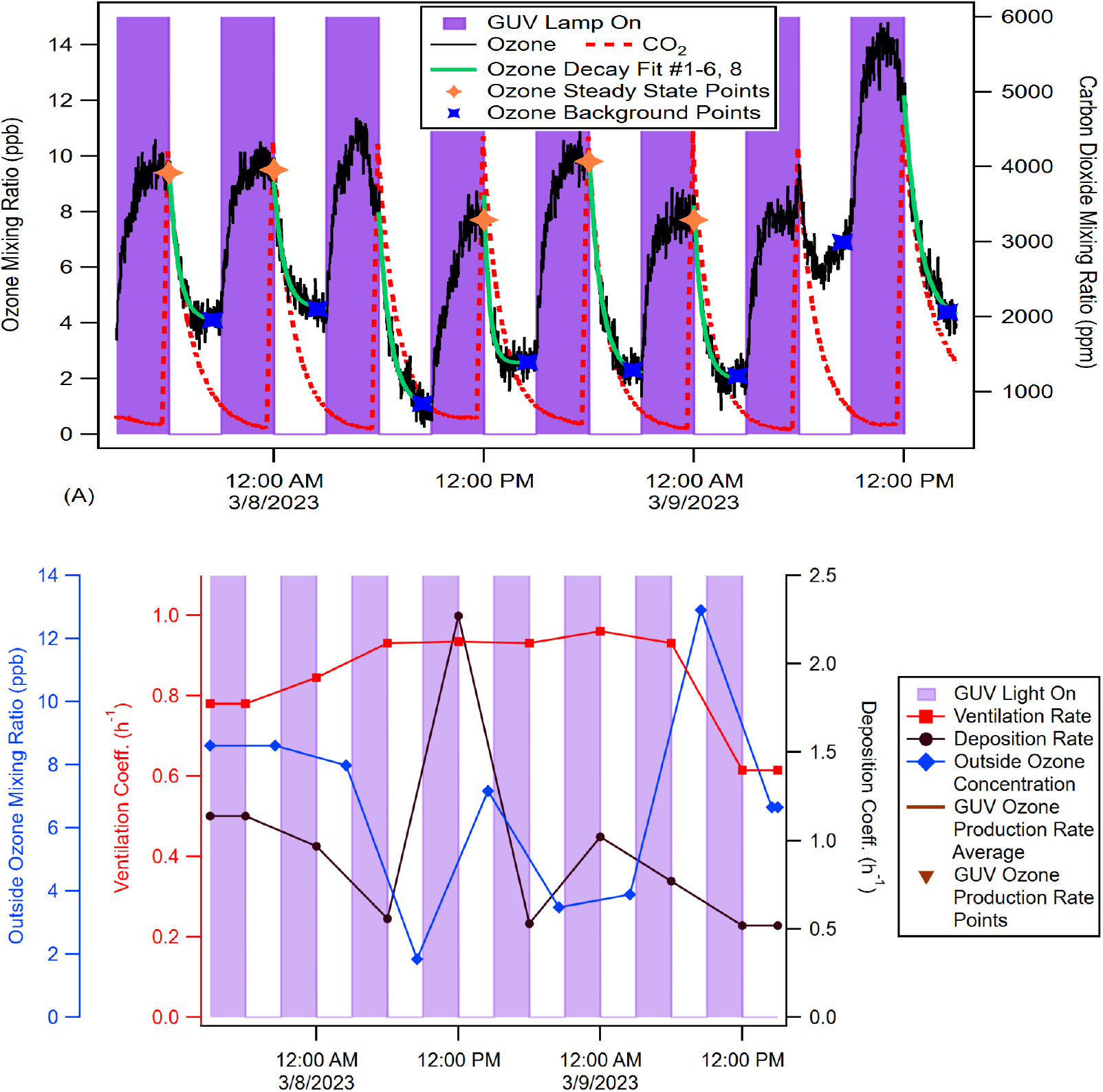
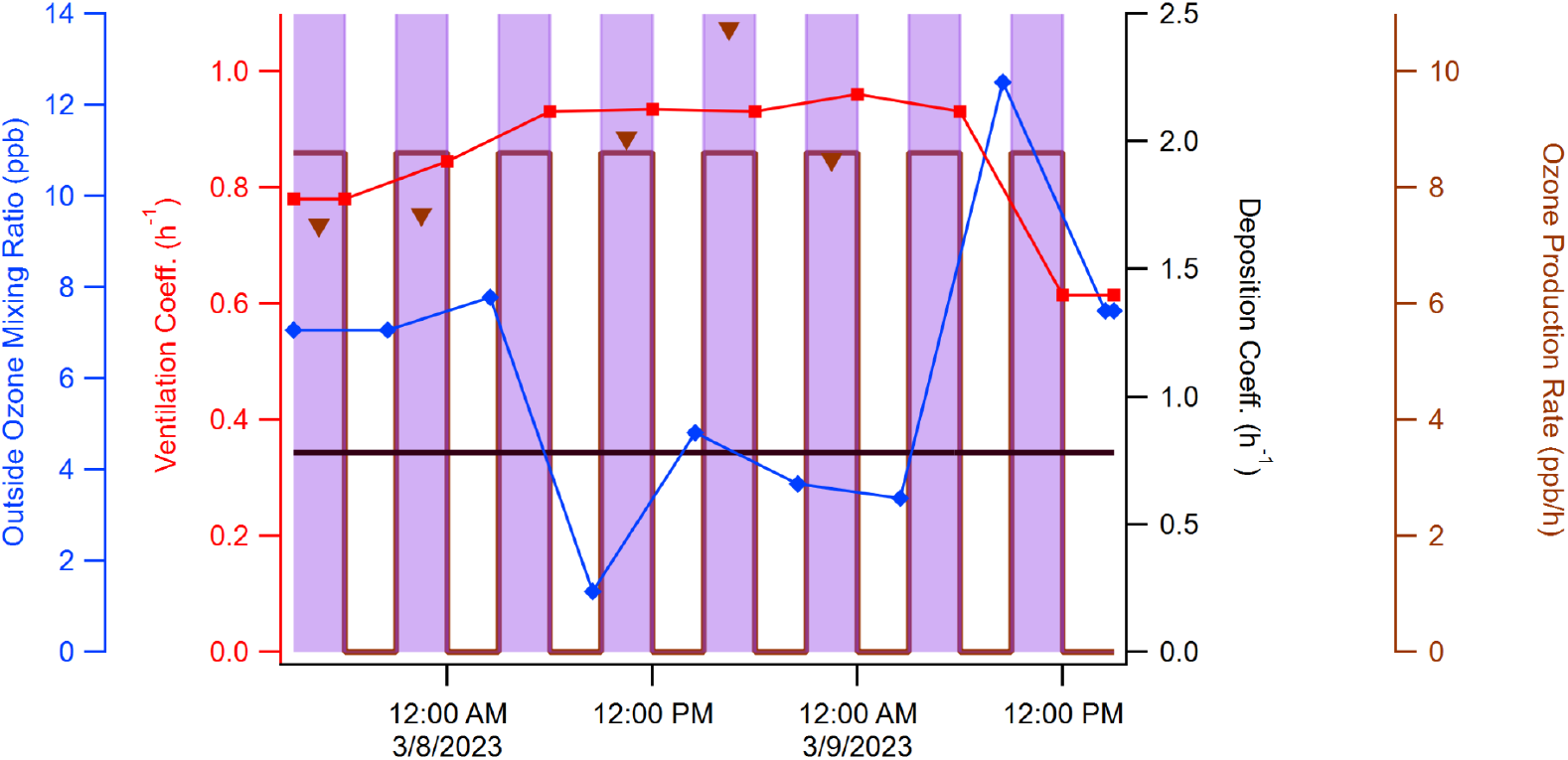
(Top): O_3_ concentration in the office as the GUV lamp was cycled on and off in 3-hour increments over 8 total cycles with corresponding CO_2_ pulses to measure ventilation rate. (Middle and Bottom): relevant parameters for modeling O_3_ concentrations in the office experiments with the KinSim model, plotted vs. time. The middle graph shows the scenario where deposition varies over time. The bottom graph shows the scenario where deposition is assumed to be constant.

**Figure S10.**
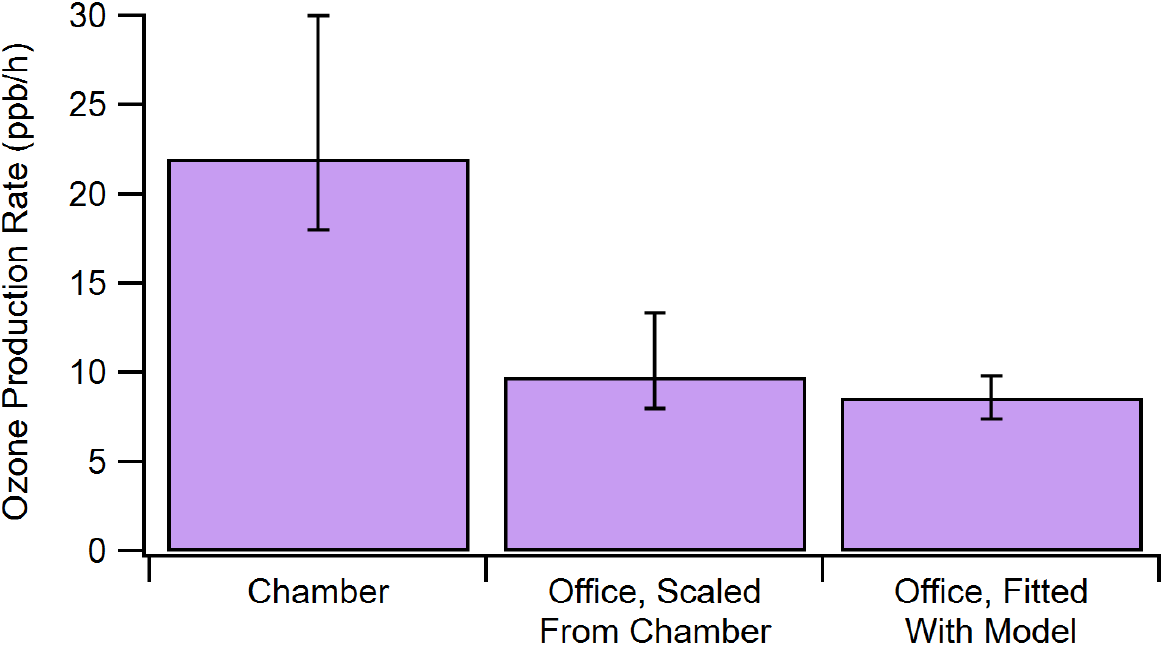
The production rates of O_3_ compared between the conference room and chamber along with the value expected for the conference room by scaling the chamber results with the relative room volume and effective GUV light pathlengths.

**Figure S11.**
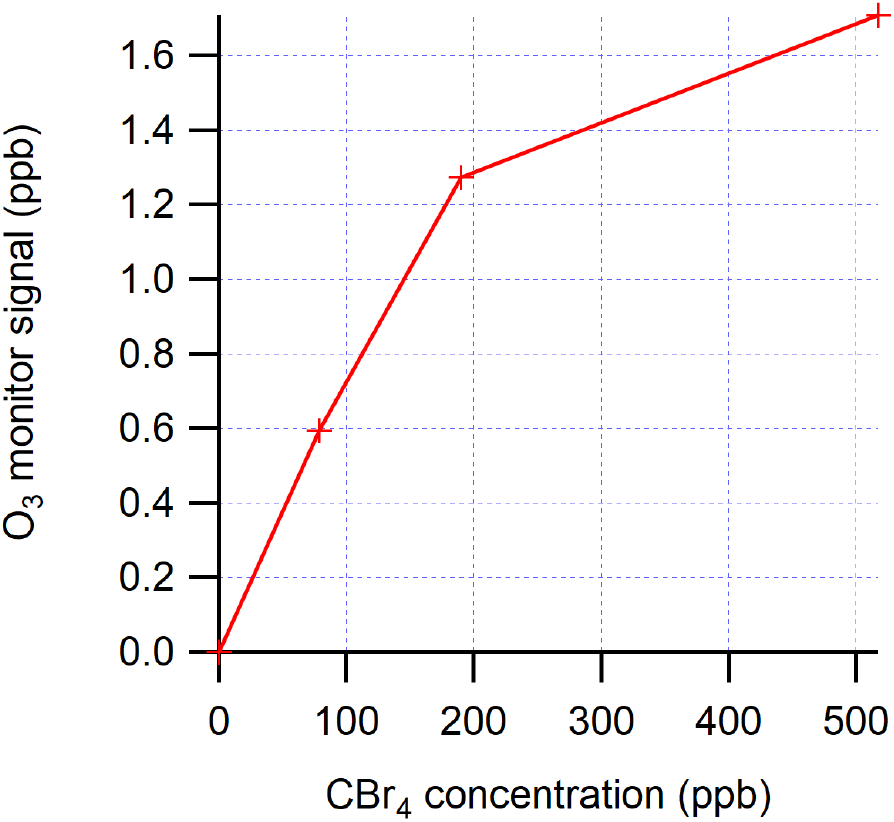
Apparent O_3_ signal measured in the chamber due to CBr_4_ interference at different CBr_4_ concentrations.

**Figure S12.**
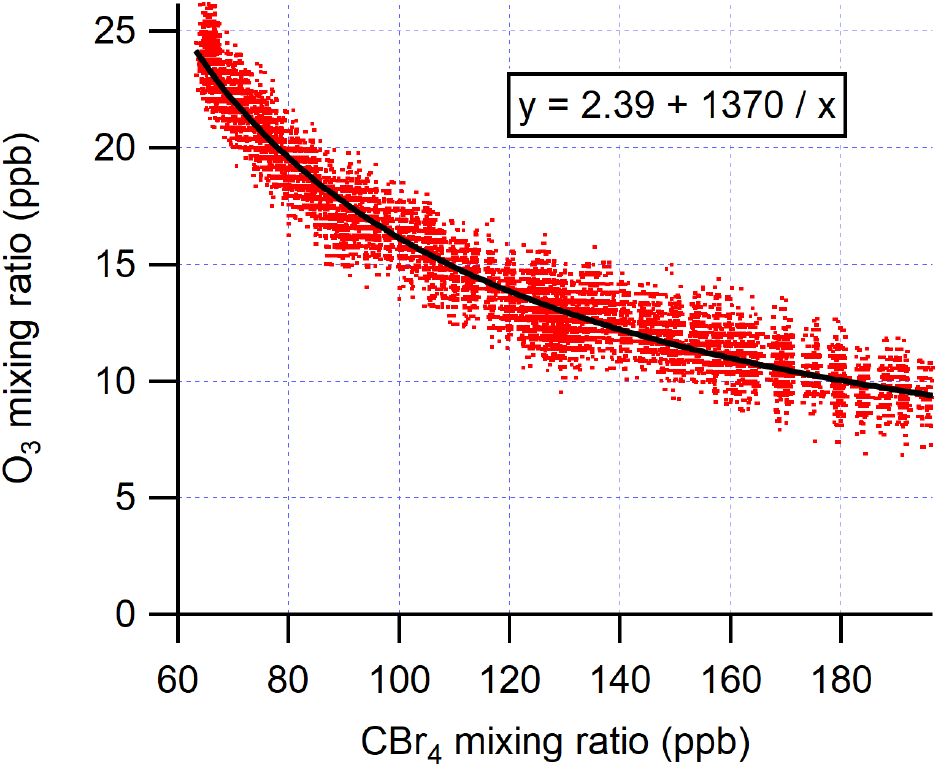
Evolution of O_3_ and CBr_4_ concentrations during a CBr_4_ photolysis experiment with the Far UV lamp (Ushio B1) lasting 12 h.

**Figure S13.**
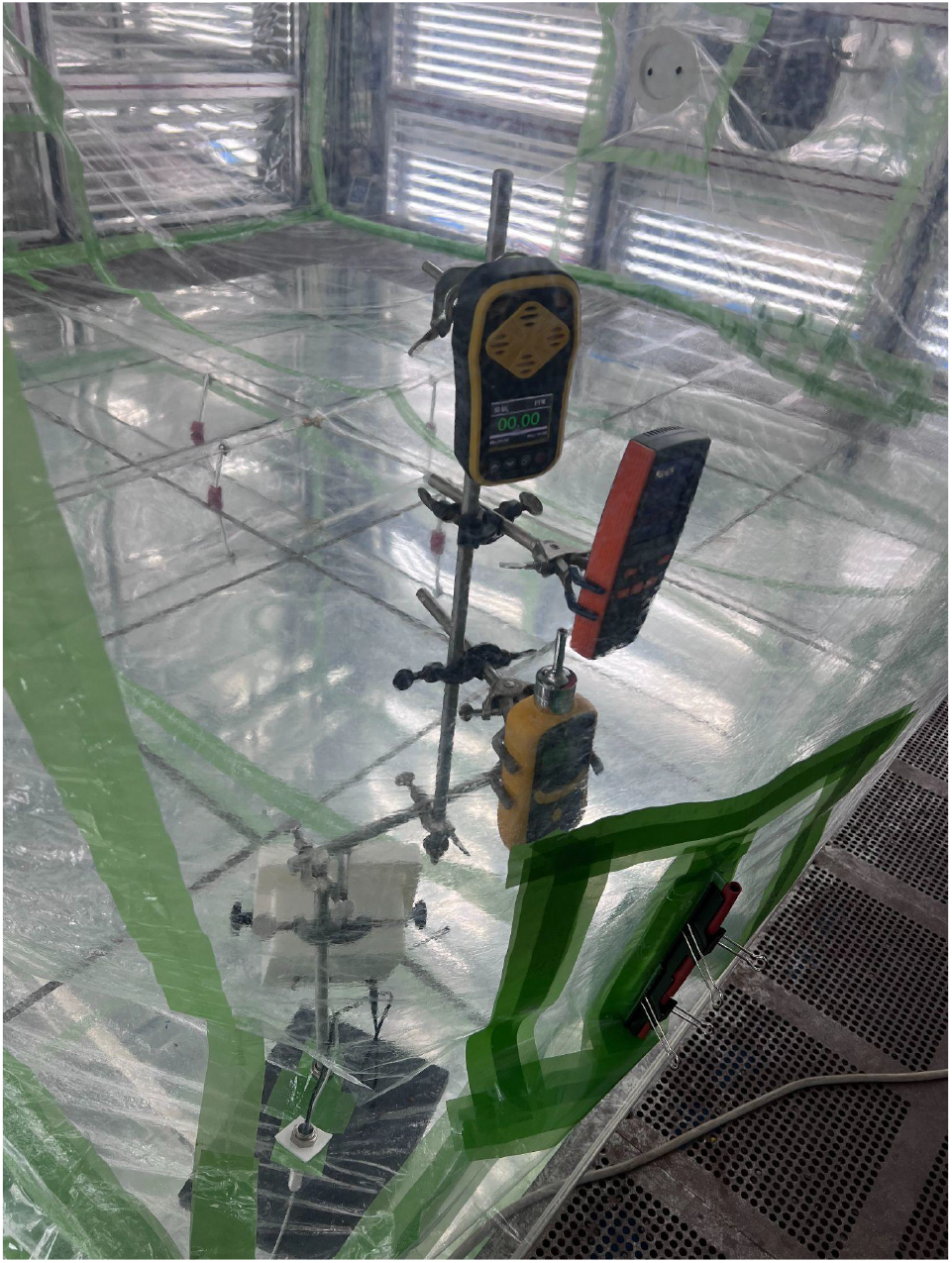
Set up with three handheld O_3_ monitors inside of the chamber. The Shandong Renke model is on top, the Shenzhen Dienmern model is in the middle, and the Shenzhen YuanTe model is on the bottom.

**Figure S14.**
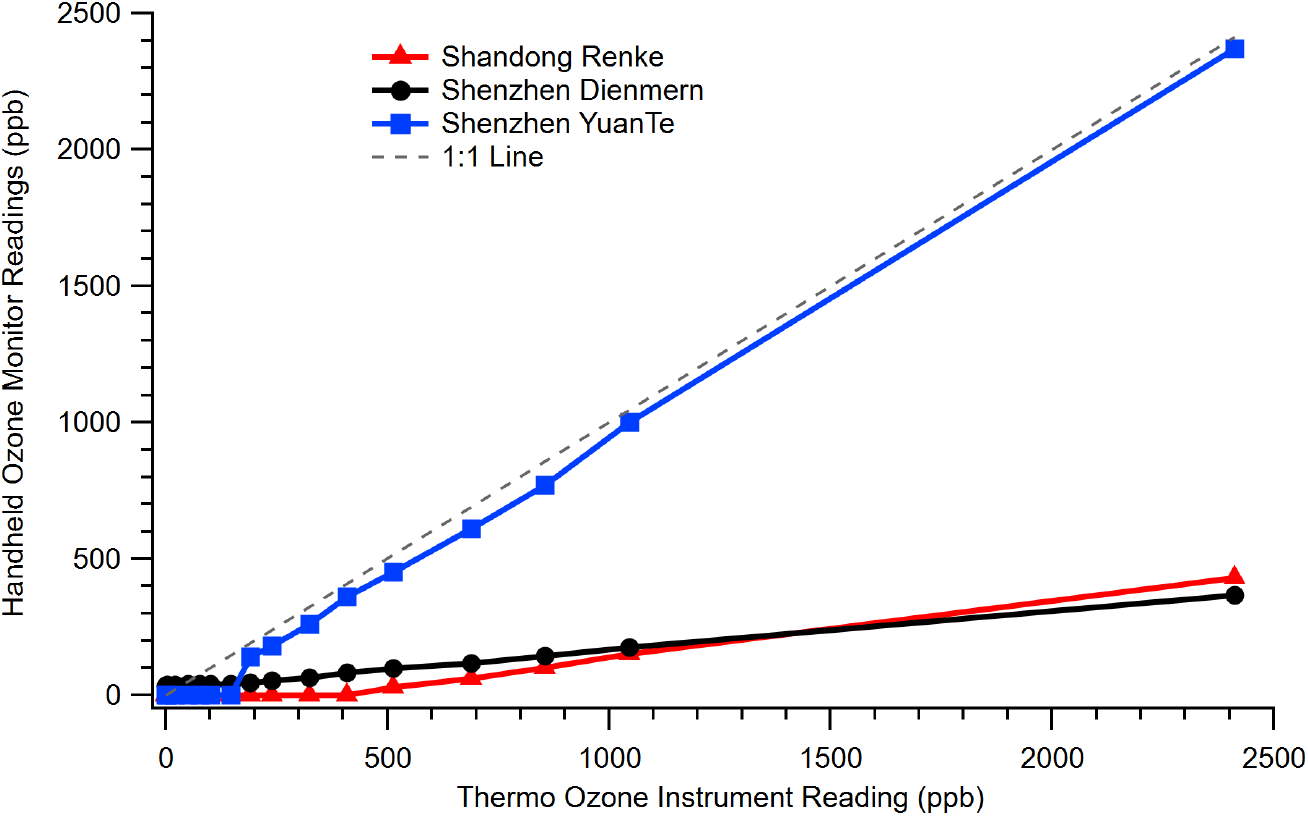
Comparison of the three portable O_3_ monitors against the research-grade Thermo O_3_ instrument.

**Figure S15.**
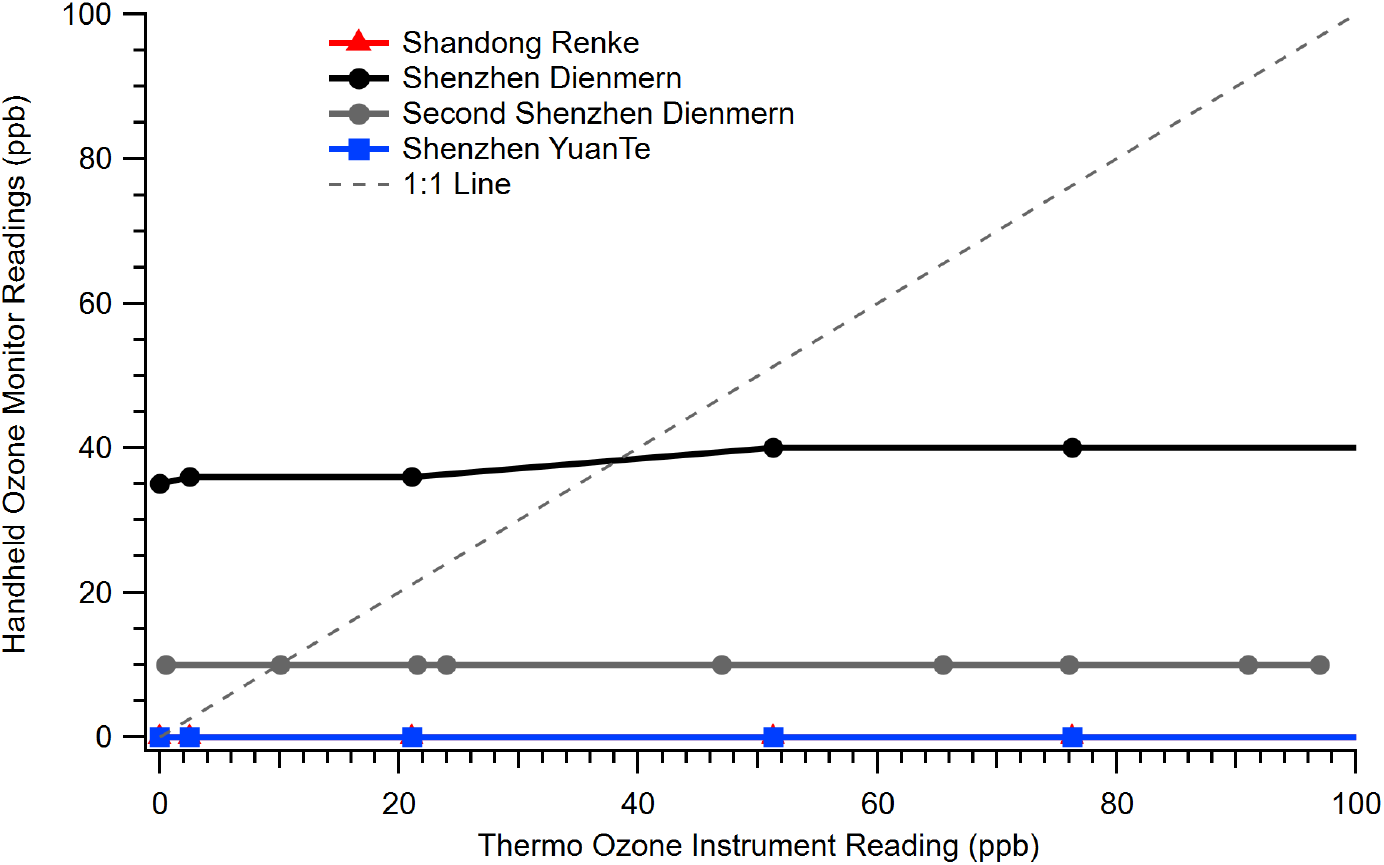
Comparison of the three portable O_3_ monitors against the research-grade Thermo O_3_ instrument, zoomed in the range of 0-100 ppb. A second and identical model Shenzhen Dienmern was also tested and shown in the gray line. This test was repeated once and none of the monitors showed appreciable differences, even the Shenzhen YuanTe still registered 0.00 ppm O_3_ (while being exposed to values in the range of this graph) after a zero calibration inside the clean O_3_-free chamber.

